# TikTok: Empowering Hidradenitis Suppurativa Patients through Social Media Education - A Cross-Sectional Content Analysis

**DOI:** 10.1101/2024.10.10.24315240

**Authors:** Paulina M. Decker, Harib H. Ezaldein

## Abstract

Hidradenitis suppurativa (HS) is a chronic condition characterized by painful, inflamed lumps under the skin in intertriginous areas. While social media platforms like TikTok are increasingly raising public awareness about HS, the quality of the information presented remains challenging to assess. Our study aims to analyze the top one hundred TikTok videos focusing on HS clinical information, treatment options, and patient experiences from 2020 to 2024. We evaluate user engagement data, content characteristics, and assess the quality of health information using a validated DISCERN score. Overall, our study suggests that while TikTok serves as a valuable resource for raising awareness and accessing new information about HS, content from healthcare professionals and scientific research garners higher user engagement and contributes to greater awareness of HS.

## Introduction

Hidradenitis Suppurativa is a chronic, recurrent skin disorder characterized by the formation of painful inflammatory nodules and abscesses, predominantly affecting flexural areas like the inguinal and axillary regions. These lesions often discharge pus and may result in hyperpigmentation and scarring.^1^ The etiology of HS involves a complex interplay of genetic and environmental factors, leading to dysbiosis of the skin microbiome and dysregulated inflammatory responses. Additionally, cardio-metabolic and autoimmune rheumatic diseases have been linked to HS due to their role in exacerbating the autoinflammatory process through the excessive release of proinflammatory cytokines.^2^ Beyond its physical manifestations, HS imposes a significant psychological and functional burden on patients, characterized by pain and malodorous discharge, further exacerbating feelings of shame and social isolation.^3^

Despite its increasing incidence, HS remains relatively underrecognized, resulting in diagnostic delays, misdiagnoses, and limited access to appropriate care. Many individuals living with HS report feeling misunderstood and marginalized.^4^ Consequently, a growing number of patients are turning to social media platforms like TikTok in search of information, support, and community. TikTok has recently emerged as a popular video sharing platform for content ranging from various topics, including medicine. Dermatological conditions have been a popular choice for content creation by dermatologist and non-medical professionals. These short videos are typically focused to create awareness about these conditions and provide advice on how to treat said conditions.^5^ Recognizing the importance of understanding the landscape of HS-related content on TikTok, our study aims to assess the quantity and quality of available evidence on the platform. Additionally, we aim to identify user preferences by analyzing patterns of engagement with educational, treatment-focused, and personal testimonial videos.

## Materials And Methods

Study Design: This cross-sectional study was conducted in February 2024. Utilizing an unbiased account created specifically for this research study, the hashtag “#hidradenitissuppurativa” was entered into the TikTok search bar and videos related to HA appeared in a randomized order.

Sample Population and Sample Size Calculation: Videos were retrieved in a randomly sorted order, with selection occurring systematically at regular intervals of three, beginning with the first video until a total of 100 videos were obtained. Exclusion criteria encompassed videos containing product promotions with commission-based incentives and those not presented in English. If a video contained any of the exclusion material listed, it was simply skipped and the following video was analyzed. Additionally, no IRB was needed due to the fact that no human subjects were needed for this study.

Video Characteristics: Various data points were collected for each video, including the number of likes, views, video duration, upload date, profile follower count, number of comments, and DISCERN score. Data collection and descriptive statistical analysis were performed using Microsoft Excel (Microsoft, Redmond, WA) in February 2024.

Content Analysis: Categorization of each video was conducted based on its content, which included general education, treatment information, and personal experiences. Videos describing HS and its signs and symptoms fell under the general education category. Videos describing different treatment methods, both medicinal and holistic, fell under the category for treatment. Lastly, videos describing an individual’s experience with HS fell under the personal experience category. Furthermore, an assessment was made regarding whether the video creator was a board-certified medical professional.

Personal Experience Videos: Videos focusing on personal experiences were scrutinized for anecdotal discussions pertaining to the patient’s symptoms, exploration of alternative medicine options, and accounts of additional procedures undergone.

Patient Education Videos and Scoring System: Irrespective of content category, each video was evaluated using the DISCERN scoring system to assess the quality of education and information pertaining to treatment choices. DISCERN comprises 15 questions, with an additional quality rating at the conclusion. Each question is graded on a 5-point scale, ranging from 1 (low quality) to 5 (high quality). This standardized tool aids in identifying bias and establishing information quality standards, facilitating consistent evaluation across all encountered HS-related videos.^6^

## Results

In this study, one hundred (n=100) TikTok videos on Hidradenitis Suppurativa (HS) were analyzed, with (n=4) 4% focusing on surgical approaches and (n=96) 96% on non-surgical methods. The videos, shared between 2020 and 2024, covered various aspects of HS, including general education, treatment options, and personal experiences. The distribution of videos peaked in 2023 (53 videos) and was lowest in 2020 (7 videos).

The analyzed videos had an average length of 41.2 seconds and a mean view count of 959,073 (Table 1). Users who posted these videos had an average of 3,197,834 followers, with videos garnering an average of 473.8 comments and 57,454 likes. The DISCERN score, which is used to evaluate the quality of health information, averaged at 2.29, with scores varying across the years. The highest mean DISCERN score of 3.09 was recorded in 2020, while the lowest score of 1.825 was observed in 2022.

**TABLE 1:** A tabulation of key data points collected for HS on TikTok in February 2024.

|  | 2020 | 2021 | 2022 | 2023 | 2024 | Total |
| --- | --- | --- | --- | --- | --- | --- |
| Video Count | 7 | 14 | 15 | 53 | 11 | 100 |
| Mean Length of Video (seconds) | 37.9 | 24.4 | 34.7 | 45.7 | 63.3 | 41.2 |
| Mean View Count | 1,414,857 | 906,800 | 698,980 | 1,040,653 | 734,075 | 959,073 |
| Mean Number of Profile Followers | 10,254,971 | 3,390,743 | 1,321,814 | 844,008 | 177,636 | 3,197,834 |
| Mean Number of Comments per Video | 875 | 448.07 | 487.7 | 295.5 | 262.9 | 473.8 |
| Mean Number of Likes per Video | 179,119 | 35,490 | 27,211 | 25,700 | 20,204 | 57,545 |
| Mean DISCERN Score | 3.09 | 2.38 | 1.825 | 1.98 | 2.19 | 2.29 |

The majority of videos (87%) originated from the United States of America (USA), with the remainder from Spain, the United Kingdom (UK), Canada, and Central America (Figure 1). U.S.-based videos exhibited significantly higher user engagement, averaging 41,888 likes and 1,046,135 views per video. The most-liked video in the United States received 959,000 likes, compared to the highest-liked videos from Spain (7,153 likes), the United Kingdom (36,500 likes), Canada (22,500 likes), and Central America received (3,573 likes).

**FIGURE 1:**
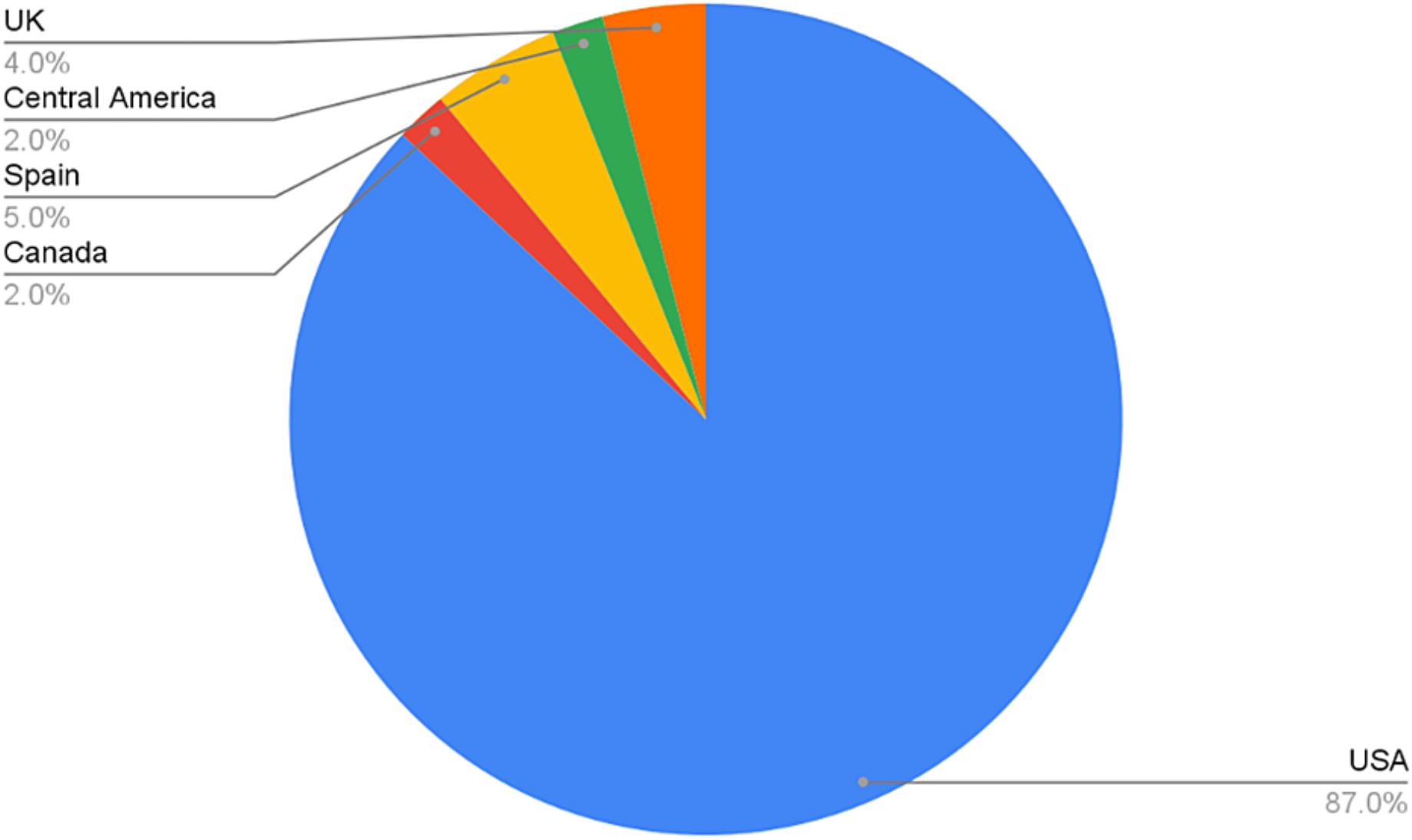
Geographic distribution of HS videos analyzed

Non-board-certified professionals produced 20% more videos than board-certified professionals (Figure 2). However, videos by board-certified professionals showed higher engagement, with an average of 53,470 likes per video. The most-liked video by a board-certified professional received 959,000 likes, while the least-liked received 123 likes. Conversely, videos by non-board-certified professionals averaged 30,662 likes, with the highest-liked video receiving 246,100 likes and the least-liked one with 19 likes. The DISCERN score for videos by board-certified professionals averaged 2.66, compared to 1.75 for non-board-certified professionals, indicating a higher quality of information provided by board-certified professionals (Figure 3).

**FIGURE 2:**
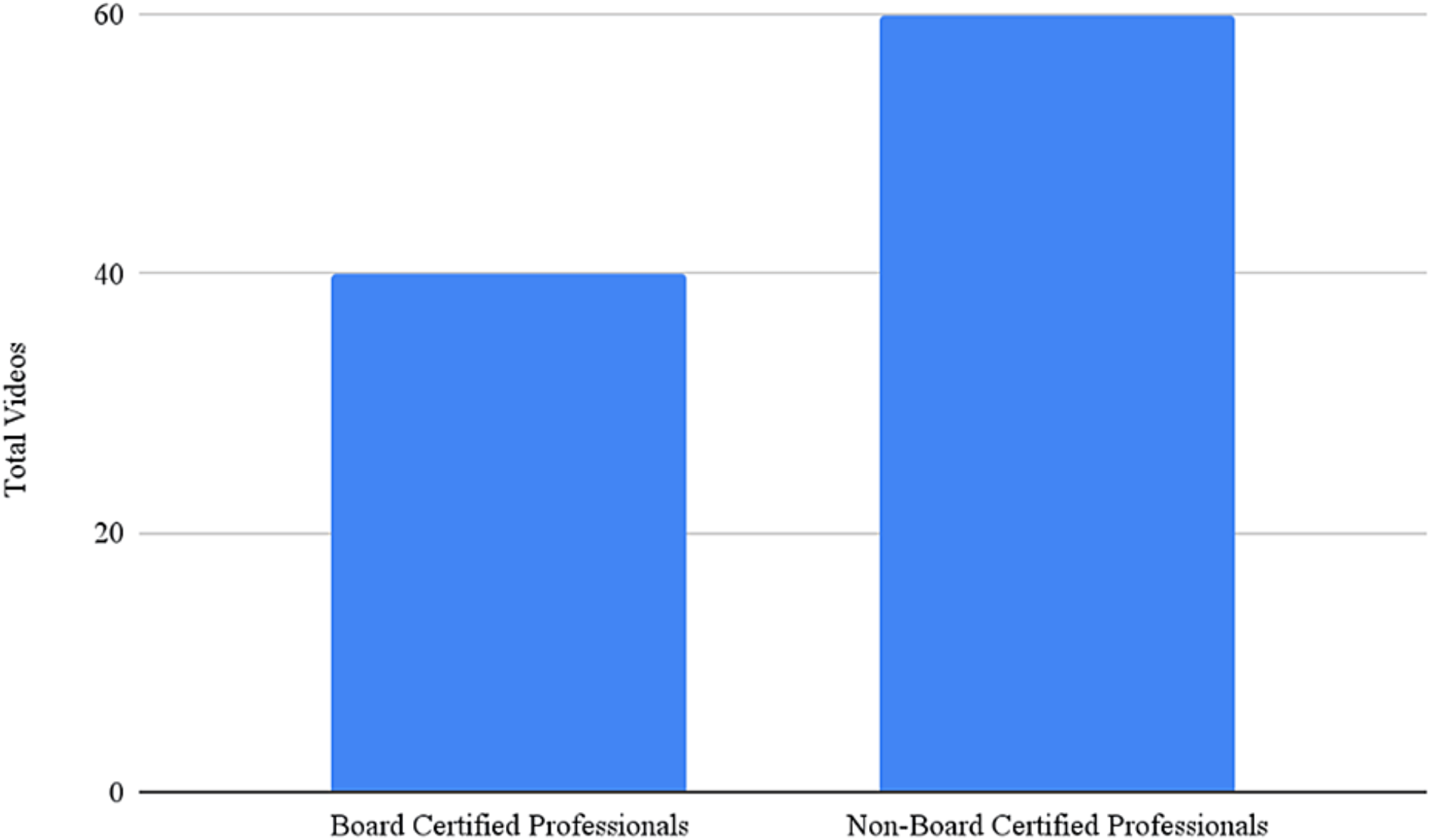
A comparison of the total number of videos between board certified professionals and non-board-certified professionals

**FIGURE 3:**
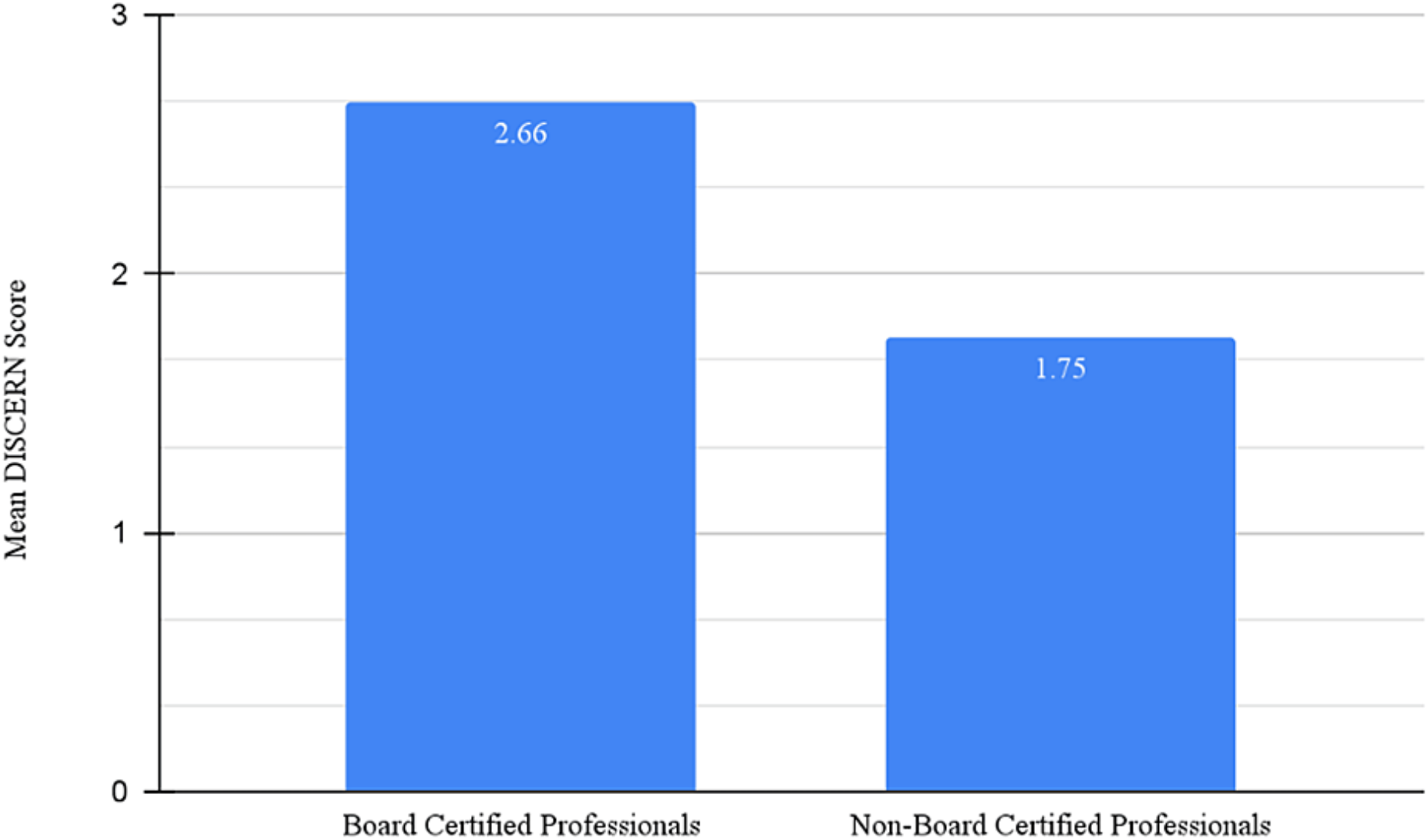
A comparison in the mean DISCERN score between videos created by board-certified professionals and non-board-certified professionals

Videos were categorized based on their content into general education, treatment, and personal experience (Figure 4). General education videos, which explained HS and its causes with minimal treatment information, accounted for 50 videos. Personal experience videos, detailing individual experiences with diagnosis, maintenance, and symptoms, comprised 32 videos. Treatment videos, focusing primarily on available treatments for HS, made up 18 videos.

**FIGURE 4:**
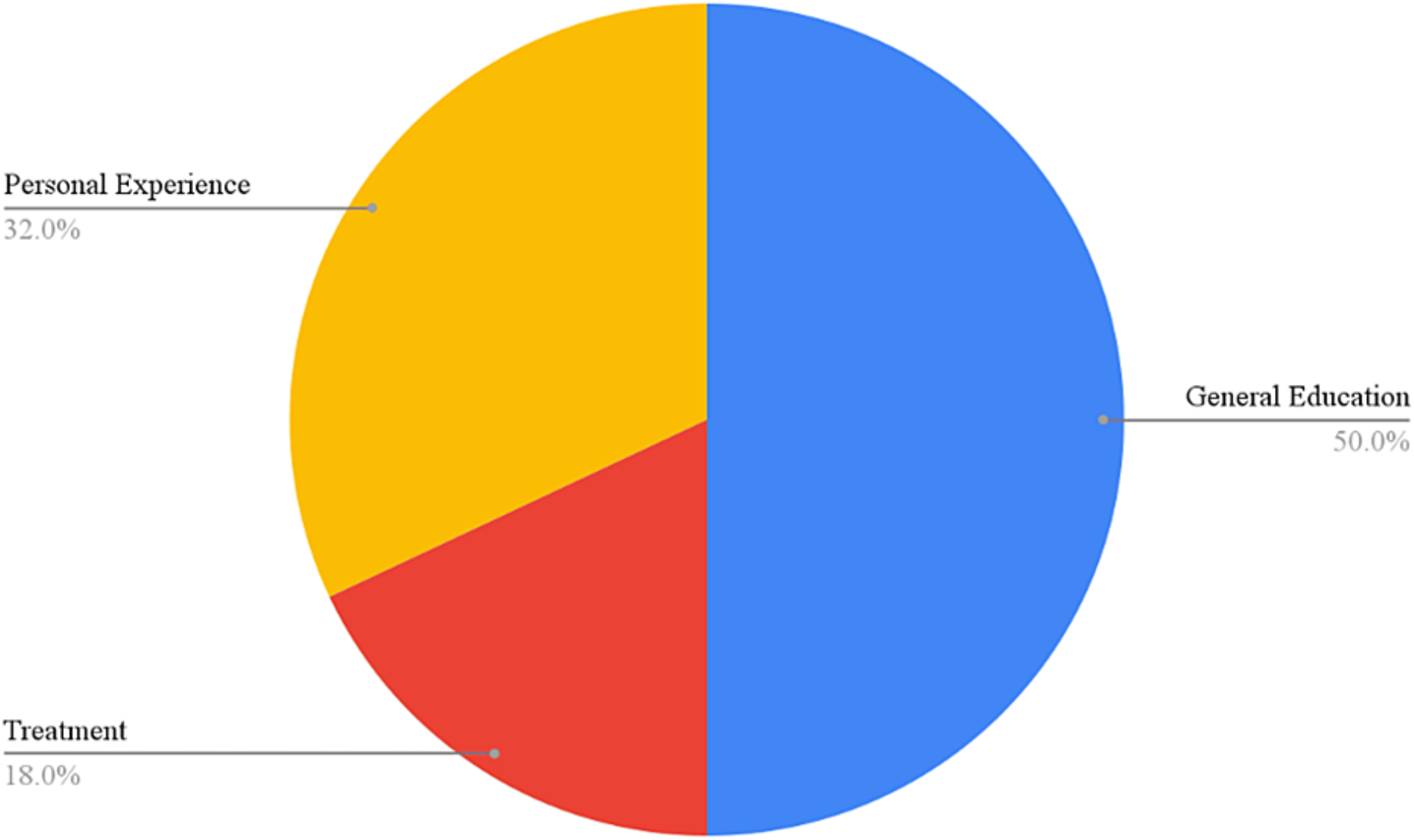
Comparison of content category for each HS video included in the study HS: Hidradenitis Suppurativa

## Discussion

This study highlights the extensive range of HS video resources available to the public on TikTok. The continual upload of new videos about HS makes it a dynamic and evolving platform for information. This is significant because it allows viewers to consistently access fresh information and personal experiences regarding HS. With over 96.3 million views and a cumulative video length of 4,280 seconds for the 100 videos analyzed, we observed notable differences in the types of information shared. These differences can be attributed to factors such as the credibility associated with board certification, the severity of HS symptoms experienced by content creators, and the creators’ willingness to openly discuss their struggles with HS.

Social media offers an accessible way for individuals, particularly younger generations, to obtain health information. TikTok has recently emerged as a leading platform for disseminating health education, albeit with some controversy regarding the validity of the information shared.^7^ Studies have shown that while social media platforms are effective in providing educational content in a timely and accessible manner and in raising awareness and support, they should not be heavily relied upon for definitive medical advice.^8^ Personal experience videos included in this study were popular due to their relatability and engagement; however, the validity of the information in these videos has not been verified through fact-checking, making them best suited as a gateway to exploring more reliable information.

A key point regarding the 32 personal experience videos analyzed is their representativeness of the HS population demographics. Of these videos, 23 creators (∼71.8%) were African American. Previous studies have reported a higher prevalence of HS among African Americans compared to Caucasians, with ratios of 1.64:1 and 1.98:1, respectively, denoting a greater incidence in African Americans.^9^ Similarly, 26 out of the 32 video creators (∼81.3%) were female, reflecting the epidemiological trend that HS is more common in females, with a reported female-to-male ratio of 3:1. Researchers attribute this gender disparity to various factors, including environmental influences, stress, hormonal differences, and genetics.^10^

The DISCERN score for videos produced by non-board certified professionals vs board certified professionals for this study have been compared to previous studies and the results support our findings. For a study completed in April of 2021 (n=100), videos by non-physicians and physicians had a mean DISCERN score of 1.63 and 2.65 respectively.^11^ Additionally, another study conducted in October of 2022, found a mean DISCERN score of 1.75 and 2.16, comparing non-board certified to board certified professionals.^12^ Our study three years later had similar findings of a non-board certified professional and board-certified professional DISCERN scores of 1.75 and 2.66 respectively. This data shows that from 2021 to 2024 the average ratio of non-physician to physician videos is consistent and roughly 1.71 to 2.49.

Several limitations in this study should be noted. The small sample size of n=100 poses a risk of not detecting differences between groups and may undermine the external validity of the findings. Focusing solely on videos in the English language limits the generalizability of the results, as many HS patients may speak other languages. Additionally, the analysis was conducted during February 2024, which could introduce a time-based limitation when comparing the number of videos and interactions per year. Newer videos might have fewer user interactions simply because they have not been available long enough, representing an avenue of future study.

## Conclusions

Hidradenitis suppurativa (HS) is becoming increasingly prevalent, making it crucial to stay informed about the latest treatment options. Social media platforms like TikTok play a significant role in raising awareness about HS, providing an accessible means for users to share information and personal experiences. For many individuals, their first encounter with HS may occur while browsing social media, indicating a growing awareness of the condition. This heightened awareness can lead to new discoveries and help reduce the stigma associated with HS. As social media continues to evolve, it will remain an essential tool in disseminating information and fostering a supportive community for those affected by HS.

## Data Availability

All data produced in the present work are contained with in the manuscript

